# Effectiveness of a nutrition education package on glycaemic control among children with type 1 diabetes mellitus aged 3-14 years in Uganda: study protocol for a cluster-randomised trial

**DOI:** 10.1101/2020.04.30.20085951

**Authors:** Nicholas Bari Ndahura, Judith Munga, Judith Kimiywe, Ezekiel Mupere

## Abstract

**Introduction:** Inadequate dietary management practices among children with type 1 diabetes mellitus (T1DM) often result in preventable complications, disability, and premature deaths, and yet strict glycaemic control can help reduce the long-term complications. Furthermore, parental caregiving has also been shown to have an impact on glycaemic control and yet often a gap exists between recommended care and provided care, resulting in failure of children with T1DM meeting their treatment targets. In Uganda, no published study has been conducted to find out if nutrition education has an effect on glycaemic control and caregivers’ level of knowledge on general and diabetes-specific nutrition for children with T1DM.

**Methods:** The study will be a cluster randomised controlled trial with 10 health facilities randomised to control or intervention at a ratio of 1:1. A total of 100 caregiver-child pairs will be recruited. The participants in the control group will continue to receive routine medical care, while those in the intervention group will receive routine medical care and a nutrition education package. The primary outcome is glycated hemoglobin (HbA1c) values. Secondary outcomes will be caregivers’ level of knowledge on general and diabetes-specific nutrition knowledge, children’s dietary diversity score and children’s mean intake of energy, protein, and fat.

**Discussion:** The findings of this study will be used in improving nutrition education in T1DM among children attending diabetes clinics in Uganda.

**Trial registration number:** The trial is registered with The Pan African Clinical Trials Registry (PACTR201902548129842).

## INTRODUCTION

Type 1 diabetes mellitus (T1DM) is one of the major forms of diabetes that affects children worldwide.^1^ T1DM is caused by the body’s autoimmune response leading to the destruction of the insulin-producing cells, therefore individuals with T1DM produce very little or no insulin. The reason why this happens is not yet fully understood but it has been attributed to several factors. ^2 3^

Prevalence and incidence of T1DM vary markedly among countries but it is estimated that 542,000 children (aged 0-14) have T1DM, with 86,000 new cases each year.^4^ T1DM is most common in Scandinavian populations such as Finland (60 per 100,000 per year) and Sweden (47 per 100,000 per year).^5 6^ T1DM is the most predominant form of diabetes in African children, ^7^ several studies in Africa reported the prevalence of T1DM as 0.33 per 1000 in Nigerian children and 0.95 per 1000 in Sudanese school children.^8 9^ In Tanzania and Sudan, incidence was reported at 1.5 per 100,000 per year and 10.1 per 100,000 per year respectively.^10 11^ A recent survey of TIDM prevalence and incidence in Rwanda reported that the prevalence of TIDM at 16.4 per 100,000 in those less than 26 years, and 4.8 per 100,000 in those less than 15 years. Incidence figures were 2.7 per 100,000 per year for those less than 26 years, and 1.2 per 100,000 per year for those less than 15 years.^12^ In Uganda, there have not been any published studies that document the incidence or prevalence of T1DM. However, recent data from 32 T1DM clinics indicates enrolment of 1187 children, this is on the increase from about 150 enrolled children in 2009.^13 14^

A Uganda health facility-based study conducted in 2018 reported more than 80% of the children having poorly controlled blood glucose levels with a mean HbA1c level of 9.7%. It was further noted that adherence to dietary recommendations was low, and most likely characterised of high intake of saturated fat and low fruit and vegetable intake. However, it was suggested that reinforcing caregiver involvement in the children’s’ diet could help improve adherence to dietary recommendations.^15 16^ In Tanzania a study reported a significant association between caregivers knowledge of diabetes and HbA1c levels.^17^ A parent-based nutrition educational intervention targeting mealtime behaviours reported a decrease in mean daily blood glucose levels among children with type 1 diabetes.^18^

Patient and parent or caregiver centred nutrition education is little explored in sub-Saharan Africa, despite being a fundamental component of diabetes education. When made easy to understand, knowledge-based, patient and parent or caregiver centred, nutrition education improves glycaemic control and helps prevent the development of complications. ^19 20^ It is therefore vital that children diagnosed with T1DM and their caregivers be educated and trained with adequate nutritional management skills and knowledge to enable them to manage and survive the onset of T1DM safely and successfully. ^21–23^ The nutritional goal for individuals with diabetes is to attain and sustain near-normal blood glucose levels by ensuring proper management of insulin therapy, physical activity, and diet. However, in children, it is important that the diet also provides for their other macro and micro-nutritional needs to ensure normal growth and development. Therefore, caregivers empowered with updated nutrition knowledge through nutrition education can serve as change agents and therefore bring about an improvement in their children’s feeding behaviour.^24^

In Uganda, there is no documented information of any study conducted to establish if nutrition education of children with T1DM and their caregivers has an effect on glycaemic control, nutrition knowledge, and dietary practices despite caregivers are crucial role to achieving optimal T1DM outcomes. Furthermore, the nutrition education module in the current diabetes education curriculum is not contextualized for the Ugandan paediatric T1DM patients and yet structured nutrition education is vital in the management of diabetes in children to empower their ability to successfully manage their condition using available resources. This, therefore, emphasizes the need to develop appropriate nutrition education interventions that are culturally and economically acceptable. ^25 26^

### Objective and hypothesis

The primary objective of the study is to implement and evaluate the effectiveness of a nutrition education package in ensuring that caregivers of children with T1DM use foods within their reach in a way that helps their children maintain good glycaemic control, ensure adequate dietary intake, diversity and improve their general nutrition knowledge.^24^ It is hypothesised that the developed nutrition education package will be more effective than the routine health education package.

### Primary outcome and measure

The primary outcome for the study is the percentage change in mean HbA1c levels of study participants. We hypothesize that children with T1DM attending clinics randomised to the intervention group receiving the nutrition education package will demonstrate significantly lower HbA1c values than those receiving the routine health education package.

### Secondary outcomes

Secondary outcomes will be caregivers’ level of knowledge of general and diabetes-specific nutrition knowledge, children’s mean intake of energy, protein, and fat, and children’s dietary diversity score. We hypothesize that caregivers of children with T1DM attending clinics randomised to the intervention group receiving the nutrition education package will demonstrate significantly improved general knowledge on nutrition in diabetes, carbohydrate counting, and food label interpretation than those receiving the routine health education package. Also, children with T1DM attending clinics randomised to the intervention group receiving the nutrition education package will have higher dietary diversity scores.

## METHODS

### Study setting

The study will be conducted at selected T1DM clinics in Uganda as indicated in Table 1. The clinics will be randomly allocated to the control or intervention group at a ratio of 1:1.

**Table 1.**
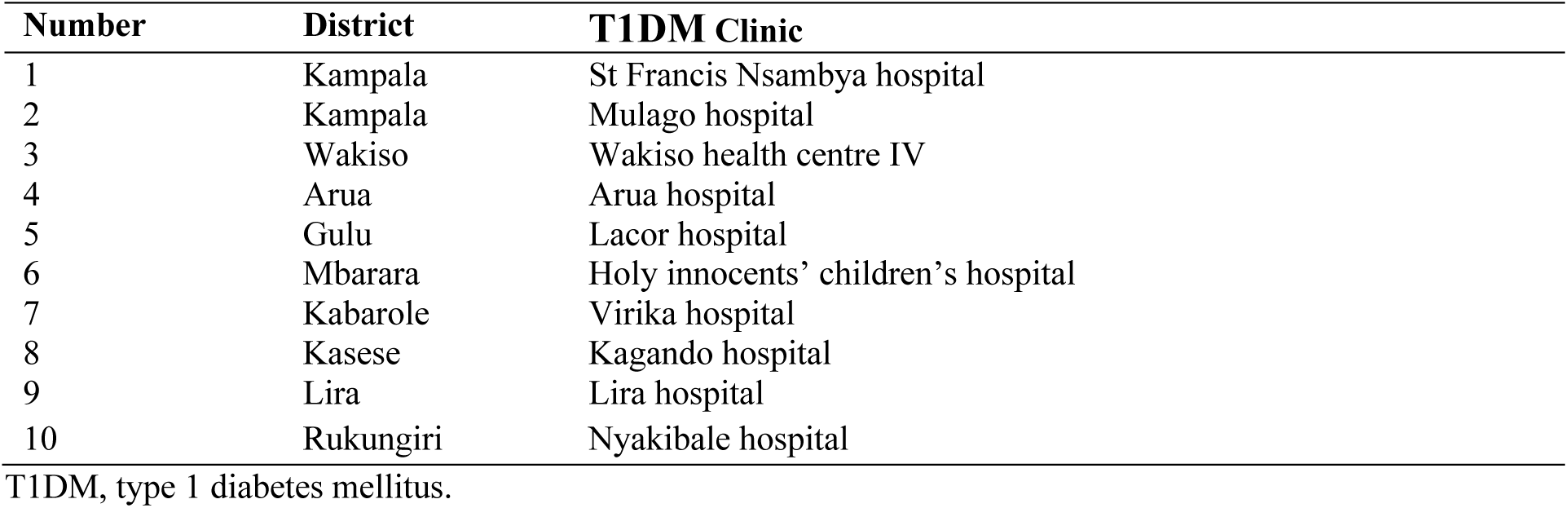
Selected diabetes clinics in Uganda

### Study design

The study design (Figure 1) will be a cluster randomised controlled trial that is SPIRIT guideline compliant.^27^ Clusters (T1DM clinics) rather than individuals will be randomly allocated to the intervention and control groups.^28^ Outcomes will be assessed at three-time points: baseline, midline, and 3-month follow-up. The control group will receive the nutrition education package after the post-intervention data collection (Table 2). A pilot study will be conducted on 20 caregiver-child pairs with similar characteristics to the sample in a diabetes clinic that will not be selected to participate in the main study. Piloting will be done particularly to validate and standardize the study procedures, research instruments and pre-test the intervention.

**Figure 1:**
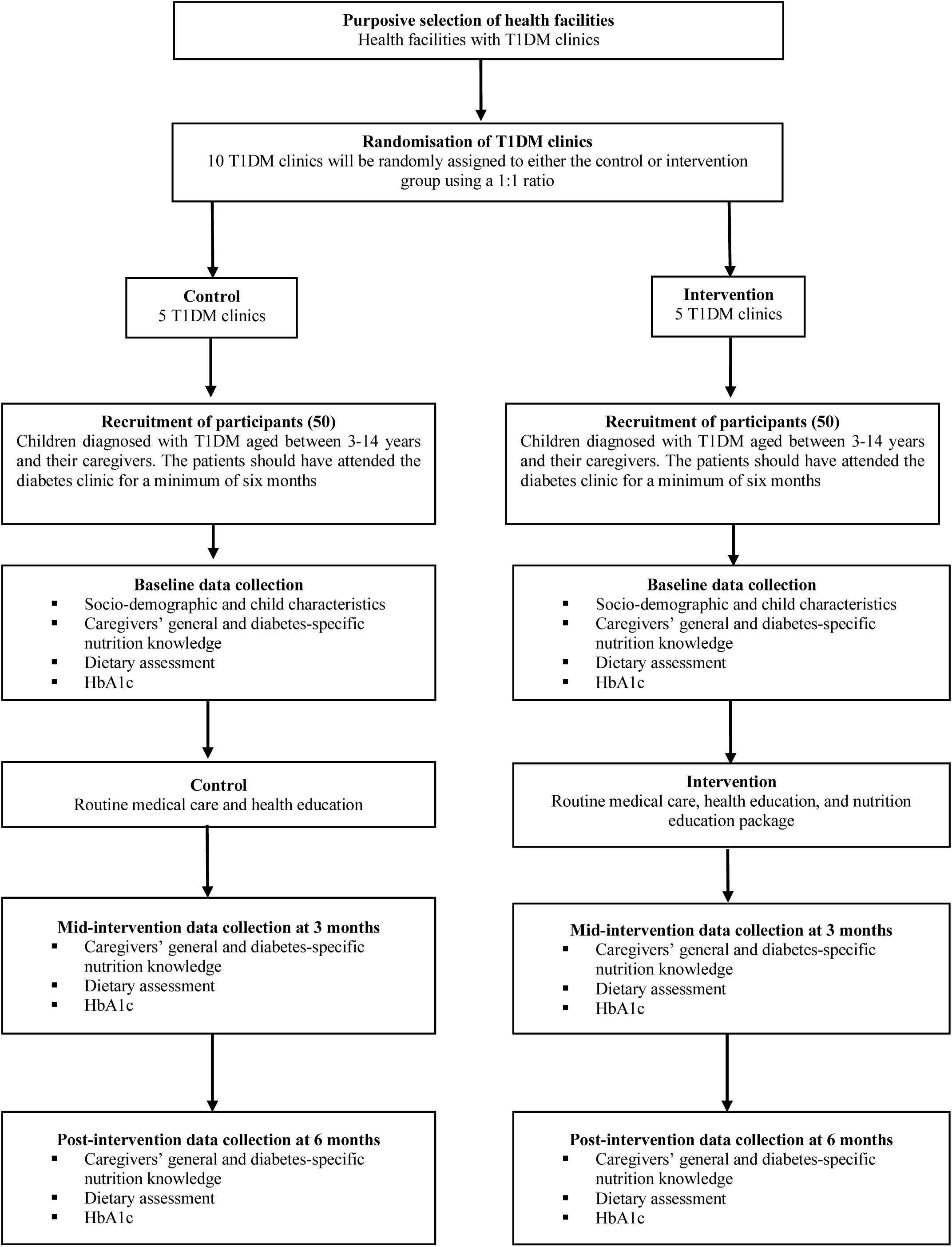
Schematic presentation of trial profile

### Study arms

The participants in the control group will continue to receive routine medical care and health education. The intervention group will also continue to receive routine medical care and health education; however, they will also attend group nutrition education sessions. Each session will last 60 minutes and will be conducted once a week for a period of 3 months. Nutrition education sessions will be conducted using food demonstrations and audio-visual aids based on the developed nutrition education guide. In addition, they will receive nutrition education materials (posters and brochures) for further reference.

**Table 2.**
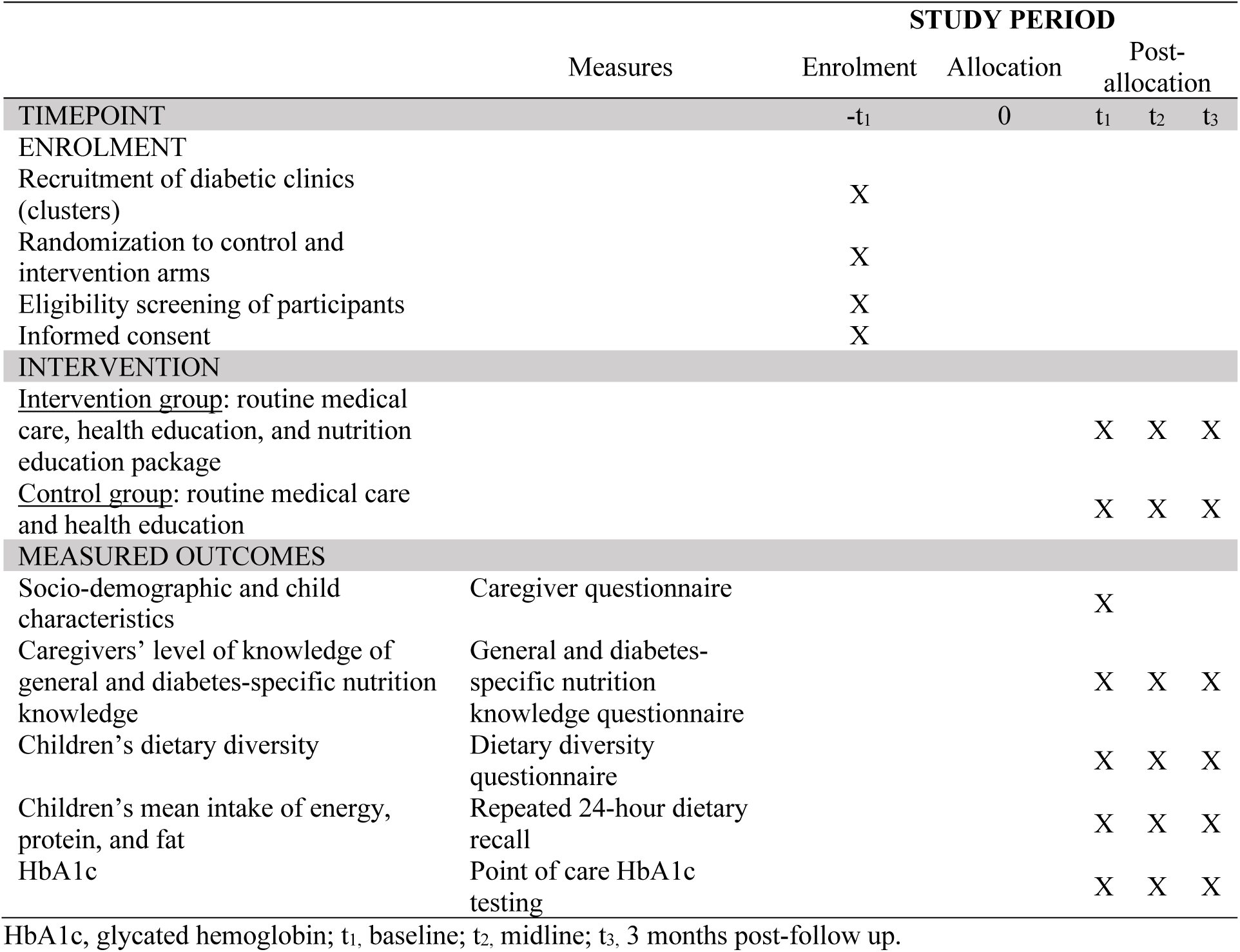
Schedule of enrolment, intervention and measured outcomes

### Sampling

A mixed-methods sampling technique will be used.^29^ 10 T1DM clinics will be purposively selected. Simple random sampling will be used to distribute the clusters into the intervention and control groups. Consecutive sampling will be used to select the study participants in each cluster due to the low number of participants in the age group (3-14 years).^30^

### Randomization

The study will comprise two study groups; a control group and an intervention group. 10 T1DM clinics (clusters) will be randomly assigned using a formula generated in Microsoft Office Excel 2016 to either intervention or control group at a ratio of 1:1. The biostatistician will be blinded to control for bias during data analysis. However, due to the nature of the intervention, the study participants will not be blinded.

### Sample size calculation

The sample size (n) was calculated using the following formula. ^31 32^

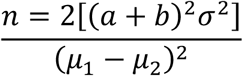

n = the sample size in each of the groups
μ1= population mean in intervention group (HbA1c = 9.9) 33
μ2 = population mean in control group (HbA1c = 8.9)
μ1-μ2 = the difference the investigator wishes to detect
σ^2^ = standard deviation (1.6)^33^
The power of the test was set at 80% and significance at 5%
a = 1.96 (conventional multiplier for alpha = 0.05)
b = 0.842 (conventional multiplier for power = 0.80, (beta = 0.20)

Therefore:

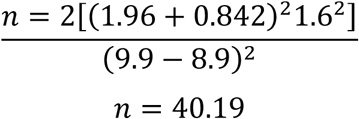

To adjust for the clustering effect the sample size was inflated by a design effect (DE) to get the adjusted sample size^34^.

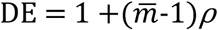

Where 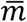 is the average cluster size as the clusters vary in size and *p* is the intra-cluster correlation coefficient (ICC). Since there is no previous study documenting the ICC, an ICC of 0.01 was considered and an average cluster size of 12 was considered.

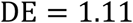

The calculated sample size was 40.19 * DE (1.11) = 44.61. The sample size will be increased by 10 *%* to 49.07 participants per study group to cater for a non-response ^35^. This was rounded off to 50 respondents in each study group, therefore a total of 100 study participants will be recruited.

### Study duration

The study is currently in the data collection phase. Recruitment for the study started in July 2019. A total of 60 caregiver-child pairs have so far been recruited. It is anticipated that mid and post-follow-up data will be finalised by December 2020.

### Recruitment of participants

Mothers/caregivers of/and children with T1DM at the diabetes clinics will be informed about the objectives of the study and the inclusion criteria. Those caregiver-child pairs that accept to participate in the study and meet the inclusion criteria will be asked to give their assent and consent respectively.

### Eligibility criteria

Mothers/caregivers of/and children diagnosed with T1DM aged between 3-14 years. The patients should have attended the diabetic clinic for a minimum of six months. In addition, the caregivers should have consented, and the children assented to taking part in the study.

### Exclusion criteria

Mothers/caregivers of/and children diagnosed with and receiving treatment for acute infections such as urinary tract infections, skin infections, respiratory infections and chronic complications such as diabetic retinopathy, nephropathy and diabetic neuropathy as illness is associated with hyperglycaemia which will impact the primary outcome of the study (HbA1c). Children who attend boarding schools will be excluded as the feeding practices are predetermined by the school management and this would introduce bias to the study.

### Intervention

The development of the nutrition education package will be guided by the following steps:

#### Step 1: Desk review of existing modules/guides on nutrition education in type 1 diabetes

A desk review of the current T1DM nutrition education module and other related material will be conducted to identify any gaps to be addressed and themes derived to guide the development of a nutrition education guide for caregivers and health care providers.

#### Step 2: Gaps in the current nutrition education module

The following gaps have been identified in the nutrition education module in the current diabetes education curriculum:

- The content is not contextualized for the Ugandan paediatric type 1 diabetes mellitus patients as examples of the foods lack more Ugandan food sources.
- Topics such as the MyPlate model, glycaemic index, the importance of fresh fruits and vegetables and how to interpret food labels are not included.
- A facilitator’s manual with structured time for delivery of every topic alongside instructional methods and teaching materials is also lacking.

#### Step 3: Setting of intervention objectives and learning outcomes

The results from steps 1 step 3 will be used to develop a matrix with competency-based objectives and learning outcomes. Relevant topics will be determined based on the objectives and learning outcomes to be achieved. Resources will be identified such as teaching aids, materials, and equipment for each topic. Time will be allocated for each topic that covers both theory and practical’s/ learning experiences. Learner-centred instructional methods will be identified for various topics.

#### Step 4: Write up of the nutrition education guide

Based on identified topics, the content will be developed, organised and put in the context of the target group. Instructor’s and participant’s manuals will then be developed.

#### Step 5: Content validation and pre-testing of the revised education package

The developed guide and facilitator guides will undergo content validation by experts from the fields of nutrition and diabetes and the target population^36^. Their evaluation will be based on four domains namely; appropriate and balanced, clarity, use of technical language and jargon and illustrations. The rejection, modification or acceptance of any content will be based on majority opinion, and all input used to improve the final draft of the guide^37^ In addition, the draft messages and Information, Education and Communication (IEC) materials will be pre-tested to determine the relevance, clarity of messages and general feeling about the messages during piloting.

### Strategies to promote adherence and completion

Individual follow-up sessions will be conducted for those individuals that may need further or any clarification on any component of the lesson. In addition, the study participants will be sent a reminder every week via the mobile phone of the next lesson and encouraged to complete all lessons.^38^ The participants will be furnished with the contacts of the researcher for any queries and clarifications.

### Primary outcome measure

HbA1c testing will be done at the different study sites using a portable point-of-care system for haemoglobin A1c testing, using the HemoCue® HbA1c 501 system (Ängelholm, Sweden). It will be calibrated as per manufacturer instructions at the time of the study. The reference levels will be < 7.5% (good control) and ≥ 7.5% (poor control). A blood sample will be obtained by pricking each child using a sterile single-use lancet. A drop of blood will then be placed at the tip of the HemoCue® HbA1c 501 patient test cartridge and the HbA1c results determined at baseline, midline and 3-month follow-up (for the intervention group only).

### Secondary outcomes measures

A validated, brief questionnaire will be used to collect information on general knowledge on nutrition in diabetes, carbohydrate counting, and food label interpretation.^39^ The repeated 24-hour dietary recall will be used to collect quantitative information on the intake of energy, protein, and fat. Caregivers will be asked to recall all the foods and fluids consumed by their children in 24 hours preceding the interview. in terms of quantities of household measures. Food photographs and volumetric vessels will be used to help the participants correctly identify and quantify the foods and drinks consumed. To reduce the random error that may arise out of the day to day food intake variation, two 24-hour dietary recalls will be conducted on a random sub-sample of 40% of the sampled caregiver-child pairs on non-consecutive days. Respondent bias will be minimized by conducting interviews on randomly selected non-consecutive days both weekdays and weekend days.^40 41^ The dietary diversity questionnaire (DDQ) will be used to collect information on the variety of foods consumed by the study participants.^42^ The DDQ will comprise of a list of different food groups from which the consumed foods from each food group will be selected based on the information provided by the respondent. Ingredients in mixed dishes in any quantity will be matched to a food group for a score of 1, each food group will only be counted once, and the total number of food groups tallied to give the dietary diversity score.^42–44^

### Other measurements

A structured questionnaire will be used to collect information on socio-demographic information (sex, age, marital status, occupation, education, religion, income, family size), and child characteristics such as insulin use, medical history, and type 1 diabetes and nutrition education training.

### Statistical analysis

The data will be analysed using IBM SPSS Statistics for Macintosh, Version 26.^45^ Descriptive summary statistics will be used to describe the characteristics of the study population and inferential statistics such as chi-square test will be used to test if there is a significant relationship between caregiver’s level of knowledge on general and diabetes-specific nutrition knowledge and the dietary diversity and adequacy of the children with T1DM. ANOVA will be used analyse the differences among group means. Paired sample t-tests will be used to determine differences between the study groups for continuous variables such as HbA1c levels, caregivers’ level of knowledge on general and diabetes-specific nutrition knowledge, mean intake of energy, protein, and fat and dietary diversity. Multiple regression analysis will be used to determine which socio, economic and demographic characteristics can predict glycaemic control (HbA1c < 7.5%). The difference in differences statistical technique will be used to test the effectiveness of the nutrition education guide in achieving glycaemic control among children with T1DM.

### Consent and participation

To ensure that study participants are as fully informed as possible about the nature of their involvement, a participant information leaflet detailing the purpose of the study, what and how the information will be obtained (procedures), withdrawal privilege, voluntary participation, and confidentiality has been developed for the children and their caregivers (supplementary file). Signed informed consent will be requested from the caregivers and the children will be asked for assent to taking part in the study after explaining to them using language that is appropriate for their age and mental capacity (supplementary file). Data of participants who withdraw will not be included in the final analysis. However, their information will be analyzed separately to determine the characteristics, reasons for drop out and lessons learnt.

### Adverse events

Caregivers shall be trained on the risks and how to recognize the early warning signs of hypoglycaemia and hyperglycaemia. And any cases of hyper or hypoglycaemia will immediately be referred to the doctor/diabetes specialist nurse for Standard Operating Procedures for the management of hypoglycaemia or hyperglycaemia. In, addition, a diabetes emergency kit/hypo box will be available at all diabetes clinics in the intervention group. The intervention will not alter the participant’s medication routine but rather will support compliance to recommendations as this will form part of the diabetes education package.

The contacts of the principle researcher, medical doctor, and diabetes specialist nurse will be provided to all participants. And they will be informed to contact them in case of an adverse event.

### Data monitoring and withdrawal

The formulation of the data monitoring team was not deemed necessary as the intervention will not alter the participant’s medication routine but rather will monitor compliance to recommendations that already form part of the diabetes education package. In addition, periodic reports will be submitted by the researcher to the REC regarding the progress of the trial and any adverse events (supplementary file). The attending clinician of the study participants has the obligation to withdraw the participant from the study should he or she anticipate that the intervention may put the participant at risk.

### Data storage

All computers with data related to the study will have the latest antivirus, anti-malware software installed, they will be password protected and kept in a secure place at all times and best practice recommendation of secure retention of data for 5 years will be adhered to. The principal investigator and biostatistician will have access to all trial data.

## DISCUSSION

The nutritional goal for individuals with type 1 diabetes is to attain and sustain near-normal blood glucose levels by ensuring proper management of insulin therapy, physical activity and dietary practices like ensuring a balance between carbohydrate intake and insulin administration. However, in children, it is vital that the diet also provides for their nutritional needs to ensure normal growth and development. A caregiver’s level of knowledge of general and diabetes-specific nutrition knowledge; in particular its nutritional management and their active involvement in their child’s diabetes management are crucial tools to achieving the above-mentioned goal.^22^ A study by Noorani, Ramaiya and Manji^17^ conducted in Tanzania reported a significant association between diabetes knowledge of caregivers with HbA1c levels. Chege and Kuria^49^ also established a significant association between dietary practices and level of nutritional knowledge among caregivers and recommended the implementation of interventions to educate caregivers on good nutritional practices. Another study conducted in Ghana also showed the tremendous effect of caregiver feeding behaviours on child nutritional outcomes.^50^ It should be noted that, most of the studies done look at the relationship between caregiver’s nutrition knowledge and its effect on a child’s nutritional status. Studies about nutrition management of T1DM among children with type 1 diabetes are limited especially in Uganda. Therefore, this study will establish the effectiveness of a nutrition education package on caregiver/child pairs on the compliance to diet and drug recommendations for attainment of normal blood glucose (HbA1c). The findings will further promote utilization of a contextualized nutrition education guide tailored to the needs of Ugandan paediatric type 1 diabetes mellitus patients at the specialized clinics and enable caregivers use foods within their reach in a way that helps their child maintain good glycaemic control and ensure adequate dietary intake. The findings will contribute to the body of knowledge on dietary management of children with type 1 diabetes mellitus.

## Data Availability

Data will be available on reasonable request.

## Acknowledgements

The authors highly acknowledge the support of the Diabetes Centre, St. Francis Hospital, Nsambya, Kampala, Uganda.

## Authors’ contributions

NNB conceived and initiated the study design. MJ, KJ, and EM contributed to the refinement of the study protocol and all authors approved the final manuscript.

## Dissemination

Results will be disseminated via peer-reviewed publications and to the health management teams at the selected health facilities. Copies will be deposited at the libraries of the participating health facilities and representative bodies for people with diabetes mellitus.

## Ethics approval

Ethics approval has been approved by the St. Francis Hospital Nsambya Review and Ethics Committee (SFHN/REC/83) and a research permit was obtained from the Uganda National Council of Science and Technology (HS186ES). Any protocol amendments will be reported to the REC and trial participants.

## Data availability statement

Data will be available on reasonable request.

## KEY MESSAGES

- This study will develop a culturally and economically acceptable nutrition education package for caregivers of children with T1DM especially in the context of Uganda.
- The study will examine the effectiveness of a nutrition education package for caregivers of children with T1DM on glycaemic control.
- The study will contribute to the exploration of nutrition education as a vital component of type 1 diabetes management in sub-Saharan Africa.

